# Youth-oriented packaging and the demand for e-liquids: Evidence from data scraped from Amazon in the United Kingdom

**DOI:** 10.1101/2021.03.13.21253514

**Authors:** Abdelaziz Lawani, Owusu-Amankwah Georgette, Ihuhwa Anna-Liisa

## Abstract

To address the threat e-cigarettes poses to public health, especially among youths, the Food and Drug Administration (FDA) issued a policy in 2020 that regulates the sale and distribution of e-cigarettes with fruit and mint flavors. Such flavors are alleged to lure youth into smoking and can increase the likelihood for addiction to other drugs. However, this regulation does not address packaging that can have a similar effect on the demand for e-cigarettes products. Indeed, certain e-liquids use youth-oriented (kiddish, cartoonish, and colorful) packaging which are attractive to youth but may also induce a no-harm perception among e-liquids users. In this paper, we examine the impact of the youth-oriented packaging on e-liquid sales. Using data scraped from Amazon, the results of our analysis reveal that youth-oriented packaging increases the sale of e-liquids. In addition, the demand for e-liquids is inelastic and the percentage of propylene glycol (PG), the rating, and the sentiment in the online reviews left by previous buyers also influence the sale of e-liquids. This research suggests that besides fruit and mint flavors, the policy goal of reducing use among youth should also include packaging. The analysis finds that taxation policies to raise prices of e-liquids will not affect appreciably the demand for e-liquids. Policies for e-liquids control should focus on designing packaging that reduces the no- or low-risk perception.

## Introduction

Introduced in the U.S. in 2007, electronic cigarettes (e-cigarettes or e-cigs), also called vapes, vaporizers, or e-pipes, refer to electronic nicotine delivery systems (ENDS) (Collins, et al., 2019). During the last decade, the demand for e-cigarettes has increased rapidly in the U.S. market. Between 2014 and 2020 total e-cigarette sales increased by 122.2% (Ali, et al., 2020). This increase in e-cigarette use, especially among the youth, is attributable to different marketing strategies such as advertising, flavors (fruit, candy, mint), and product types (prefilled vs disposable products) (Ali, et al., 2020). These marketing strategies are associated with a non-harm perception and intention to use e-cigarettes and pose a public health hazard if unregulated (Collins, et al., 2019).

E-cigarettes are generally marketed as safer alternatives to traditional smoking and as aids to reducing or ending smoking (Grace, et al., 2014, Tucker, et al., 2017). However, recent evidence suggests e-cigarettes pose public health risks (Eaton, et al., 2018, Etter, 2018) since they can serve as a gateway into smoking and may increase the long-term dependency on tobacco products (Schneider and Diehl, 2016). E-cigarettes can also reduce the use of clinically-proven smoking cessation products among those who want to quit (Tuchman, 2019). They can cause considerable harm to the population of youth because of their “safe” impression. Youths that use e-cigarettes in nontraditional flavors (for example fruit and candy) are more likely to continue vaping (Leventhal, et al., 2019). In January 2020, 82% of hospitalization in the U.S with an e-cigarette or vaping product use–associated lung injury (EVALI) are strongly linked to the utilization of smoking devices (Krishnasamy, et al., 2020). Examining the bronchoalveolar-lavage (BAL) fluid from 51 patients with EVALI, Blount, et al. (2020) confirm that injury in the lung is associated with the use of e-cigarettes.

Policy makers have set regulations to control the demand for e-cigarettes, especially among youth, that range from minimum legal sale age laws to excise taxes on e-cigarettes (Pesko, et al., 2018). In 2020, the Food and Drug Administration (FDA) issued an enforcement policy against the manufacture, distribution, and sale of unauthorized flavored prefilled pods or cartridge-based e-cigarettes, including fruit and mint flavors that appeal to youth (Wang, et al., 2020). However, similar to fruit and mint flavors, packaging can also lure teens into smoking. Indeed, youth-oriented attractive, colorful, and cartoonish packaging can produce a low-harm perception among youth, enticing them into e-cigarettes consumption. However, there is little scientific evidence on the effect of packaging on the demand for e-cigarettes.

The present study fills this gap by examining the effect of packaging on the demand for e-liquids or e-juices which are liquids aerosolized by the e-cigarettes. A large and growing proportion of e-cigarette and e-liquids being sold on online platforms (Ali, et al., 2020) where the vast majority of youth that use these products are active we also examine the effect of reviews shared on online platforms on sales. We collected data on e-liquids sold on Amazon in the United Kingdom and find that e-liquids with youth-oriented packaging have a greater influence on the product’s sale rank. In addition, online reviews, and ratings, also explain the demand for e-liquids. However, the demand for e-liquids is not price elastic, suggesting that policies that aim at increasing e-liquids prices through taxation will not be effective in reducing e-liquids use.

The rest of the paper is structured as follows. The second section discusses related work. In section 3, the data and the empirical estimation strategy for our analysis are presented. The results of the econometric analysis are discussed in section 4 and section 5 concludes.

### Related literature

In recent years, the purchase and use of electronic cigarettes (e-cigarettes) among youth across high-income countries has increased at alarming rates. E-cigarettes are making nicotine use among the youth a regular activity (Walley, et al., 2019). Hammond, et al. (2020) finds e-cigarettes to be the most prevalent nicotine product among US youth, with 14.5% of them reporting frequent use in 2017. The rate of e-cigarette use among the youth population surpassed adults in 2014 (McMillen, et al., 2015). Similar increasing trends have been observed among high-income countries including youth in Canada (Czoli, et al., 2014, Reid, et al., 2015), China (Xiao, et al., 2018), Saudi Arabia (Awan, 2016), South Korean youth (Lee, et al., 2017), the UK (Conner, et al., 2018) and 27 European Union member States (Filippidis, et al., 2017).

The appeal of e-cigarettes to youth poses a public health threat. Among the different factors that lure youth into smoking, Ali, et al. (2020) identify the different flavors such as fruit, candy, and mint that are attractive to youth. Although flavorings in food and drinks are perceived as safe to be ingested, flavoring in e-cigarettes may not be safe to inhale and e‐cigarettes emit considerable levels of toxicants that are harmful to vulnerable populations (Lu, et al., 2020). E-cigarette companies commonly advertise that e-cigarettes contain nicotine, flavoring chemicals, and humectants (propylene glycol and/or vegetable glycerin), but toxicants, ultrafine particles, and carcinogens have also been found in e-cigarette solutions and emissions. Many of these later chemicals are known to cause adverse health effects (Walley, et al., 2019). E-cigarettes primarily affect the pulmonary system but also have effects on the immune system, the cardiovascular system, and the central nervous system (MacDonald and Middlekauff, 2019, Qasim, et al., 2017). E-cigarette aerosols contain particulate matter and metals harmful to human health (Eaton, et al., 2018),

The variety of e-cigarettes flavors available to youth can increase their consumption and therefore exposure to negative health impacts. According to Walley, et al. (2019), there are an estimated 15,000 e-cigarette flavors, including products with labels enticing to children and adolescents that imitate cookies, whipped cream, alcoholic beverages, and other dessert flavors. Marketing strategies such as video advertisements on television and other electronic platforms used by e-cigarette companies have been the most pervasive and are associated with an uptick in use among youth (Struik, et al., 2020). Marynak, et al. (2018) estimate that 20.5 million U.S youth had been exposed to at least one source of e-cigarette advertisement in 2016.Youth that are exposed to advertisements are more likely to experiment with (Chen-Sankey, et al., 2019) and use e-cigarettes (Mantey, et al., 2016).

Regulations have addressed marketing strategies such as advertisement for tobacco products since 1971 but not e-cigarettes (Tuchman, 2019). Regulations have a significant effect on how vendors sell nicotine-based vaping products to the youth but may not adequately achieve goals of protecting the health of youth through restricted access (Kilcommons, et al., 2020). One challenge related to the regulation of e-cigarettes is the variety and number of devices and flavors used in e-cigarettes. For example, Lanza, et al. (2020) examine the challenges related to the optimality of regulations targeting the wide spectrum of vaping products in protecting the health of the youth. Stressing the limit of federal regulations in protecting youth from e-cigarette use, exposure, and nicotine addiction, Walley, et al. (2019) recommend to pediatric health care providers to counsel and advocate for a tobacco-free lifestyle among the youth. In the U.S., the Food and Drug Administration (FDA) has developed a set of policies that regulate e-cigarettes flavors such as fruit and mint that appeal to youth. However, even though packaging has been shown to influence consumers’ perception of a product and their purchasing decision (Hamdar, et al., 2018, Hussain, et al., 2011, Muhammad and Kamran, 2014, Raheem, et al., 2014), these policies do not include regulations on packaging attractive to youth that can also induce the use of e-cigarettes. For example, Venter, et al. (2011) show that food packaging provides prospective consumers with virtual stimuli necessary to gain their attention and form an opinion on the product quality. Fraser (2018) associates aesthetic food packaging with the purchasing intent of consumers. Al‐Samarraie, et al. (2019) finds that packaging contents such as colors, graphics, label information, and country of origin were the main determinants that influence consumers’ decision to buy. However, in the literature related to e-cigarettes, there is not enough evidence on the impact of packaging on purchases. The present study addresses this issue by examining the effect of packaging on the sale of e-liquids. It also discusses the effect of online platforms on the demand for e-liquids.

Analyzing online platform trends is a meaningful way to inform public health practitioners of current sentiments regarding e-cigarettes (Cole-Lewis, et al., 2015). Internet advertisement exposure has been associated with lower perceived harm of e-cigarettes (Reinhold, et al., 2017). Besides social networks, social acceptance, product-related - availability of flavors cited as reasons for e-cigarette use among youth (Struik, et al., 2020), an important motivation is the perception that they are less harmful than cigarettes (Kinouani, et al., 2017). College tobacco users perceive e-cigarettes to be healthier and cleaner than traditional cigarettes (Kong, et al., 2015). Kwon and Park (2020) examine the sentiments associated with e-cigarettes on online platforms. Their results indicate that although perceptions among social media users are mixed, positive sentiments were more often expressed than negative perceptions. Lu, et al. (2020) also confirm the significant positive sentiments most e-liquids have on online platforms. Online platforms is dominated by pro-vaping messages disseminated by the vaping industry and vaping proponents, while the uncertainty surrounding e-cigarette regulation expressed within the public health field appears to be reflected in ongoing social media dialogues (McCausland, et al., 2019).

## Econometric model and data

### Econometric Model Specification

We model the effect of packaging on the demand for e-liquid by adapting a two-level mixed-effects linear regression model (Forman, et al. (2008), Ghose and Ipeirotis (2010), Hu, et al. (2014) and Yang, et al. (2016)). The mixed-effect linear model offers the advantage to incorporate a hierarchical structure for the dataset and account for brand and e-liquid specific factors that influence the demand for e-liquids that have not been incorporated in the model. The mixed-effect linear model also creates unbiased estimates (Orford, 2000, Yang, et al., 2016).

The model is specified as follows:

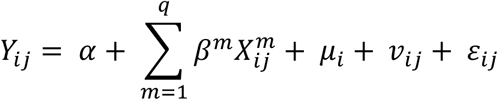

where *j* indicates the e-liquid (the first level of observation) and *i* indicates the brand (the second level) to which the first level observation belongs. *Y* is the dependent variable. It represents the sales rank as in Ghose and Ipeirotis (2010). According to past literature, the sales rank can be used as a proxy for the demand for products on Amazon (Chevalier and Mayzlin, 2006, Forman, et al., 2008). Chevalier and Goolsbee (2003) show that there the observed sales rank of a product on Amazon is correlated to its unobserved demand. Products that sell well on the online platform have a better rank (lower number). This inverse relationship between sales rank and demand influences the interpretation of the estimate from the econometric model. We use the log of sales rank to address non-linearities and smooth large values. *α* is the constant term. We have *q* explanatory variables represented by *X*. Based on past literature and data availability, we included product-specific characteristics and online reviews that have also been shown to influence purchasing decisions on Amazon (Chevalier and Mayzlin, 2006). We use as explanatory variables for each e-liquid, the selling price of the e-liquid in pound sterling, the volume of the e-liquid in milliliters, the average rating score, the average number of reviews, the average length of the reviews, the average sentiment score of the reviews, a dummy variable that represents e-liquid designed for all types of vapers, a dummy variable identifying e-liquids that have a youth-oriented packaging which is our main variable of interest, and the concentration in Propylene Glycol.

The *μ*_*i*_ denote brand-level random effects and captures unobservable heterogeneities across brands. *v*_*ij*_ denotes the e-liquid-level random effects that captures unobservable characteristics of the e-liquids. The error term is represented by *ε*_*ij*_. The random effects *μ*_*i*_ and *v*_*ij*_ and the error term *ε*_*ij*_ follow an independent normal distribution with mean 0 and unspecified variance.

### Data

We scrapped data on e-liquid (e-juice or vape juice)^1^ sales on Amazon in the United Kingdom. Amazon is an online platform that connects buyers and sellers of goods. The e-liquid is the liquid transformed by an e-cigarette into an aerosol. It is often a mix of water, food-grade flavoring (tobacco, mint, menthol, etc.), propylene glycol (PG), vegetable glycerin (VG), and different levels of nicotine, or other cannabinoids extracts. The presence and concentration of the different components differ from an e-liquid to another. Sales quantities are not available through Amazon, but sales rank is. Product-specific characteristics such as retail price, packaging, and sales rank (used as a proxy for sales) were collected. We also collected data on the volume of the e-liquid, whether or not it was a short-fill drip, and the ratio PG/VG. Short-fill e-liquids are designed to offer flexibility to e-cigarettes users to adjust the nicotine concentration. Bottles of e-liquids are not fully filled but space is left so that the user can add and mix the e-liquid with nicotine. For instance, a 60 ml short-fill e-liquid can have a bottle filled with 50ml of juice leaving room for the end-user to add up to 10 ml of nicotine. Short-fill e-liquids offers more choices to end users since they can adjust the dosage of nicotine. PG and VG are the two primary solvents of e-cigarettes and the ratio PG/VG determines the consistency and intensity of throat hit of the e-liquids. Higher concentrations of PG produce a better throat hit while higher concentrations of VG produce less throat hit but more vapor. However, the composition of PG and VG in e-liquid has been shown to damage lung function even with no nicotine (El-Hage, et al., 2020, Madison, et al., 2019). To determine the type of packaging, we consulted the listing of each e-liquid in our sample on Amazon and manually coded them as “youth-oriented” and “non-youth-oriented”.

**Figure 1:**
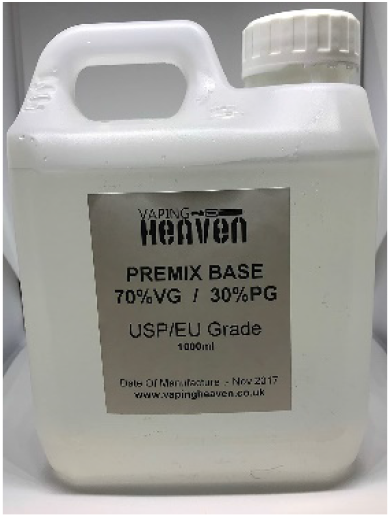
Example of e-liquid with non-youth-oriented packaging

**Figure 2:**
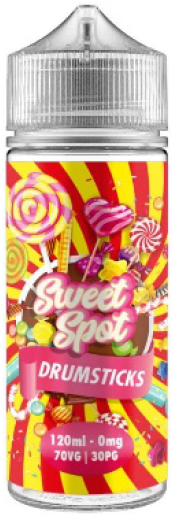
Example of e-liquid with youth-oriented packaging

On online platforms such as Amazon, buyers can also leave a review on the goods purchased. The reviews can affect the purchase decision of other buyers (Forman, et al., 2008, Hu, et al., 2014) and in the case of e-liquids, the sentiments in the reviews, as well as the specific attributes discussed, can inform prospective buyers on the product quality and facilitate their decision to buy the product (Cole-Lewis, et al., 2015, Lu, et al., 2020). We collected data on the number of reviews, the text in the review as well as the review rating. Besides numeric ratings, the sentiment conveyed in product reviews has been shown to affect consumer purchasing decisions (Guo, et al., 2012, Hu, et al., 2014). To account for the effect of the sentiment in reviews on sales we follow (Lawani, et al., 2019) to detect and score positive and negative sentiment in the e-liquid reviews. We removed punctuations, numbers, non-textual contents, and irrelevant words from the reviews. We then extract the root of the word or stem from the remaining words in the reviews and each stem is attributed a sentiment score based on its match in the AFINN lexicon. The AFINN lexicon is developed by Nielsen (2011) and comports English words with their associated sentiment score. The score of each review in our study is the sum of the score of the words used to write the review.

A total of 307 products categories were collected on Amazon. After cleaning the data from unrelated products, 139 e-liquids are analyzed in our dataset.

Table 1 gives the description and summary statistics of the main variables used in our study.

**Table 1:**
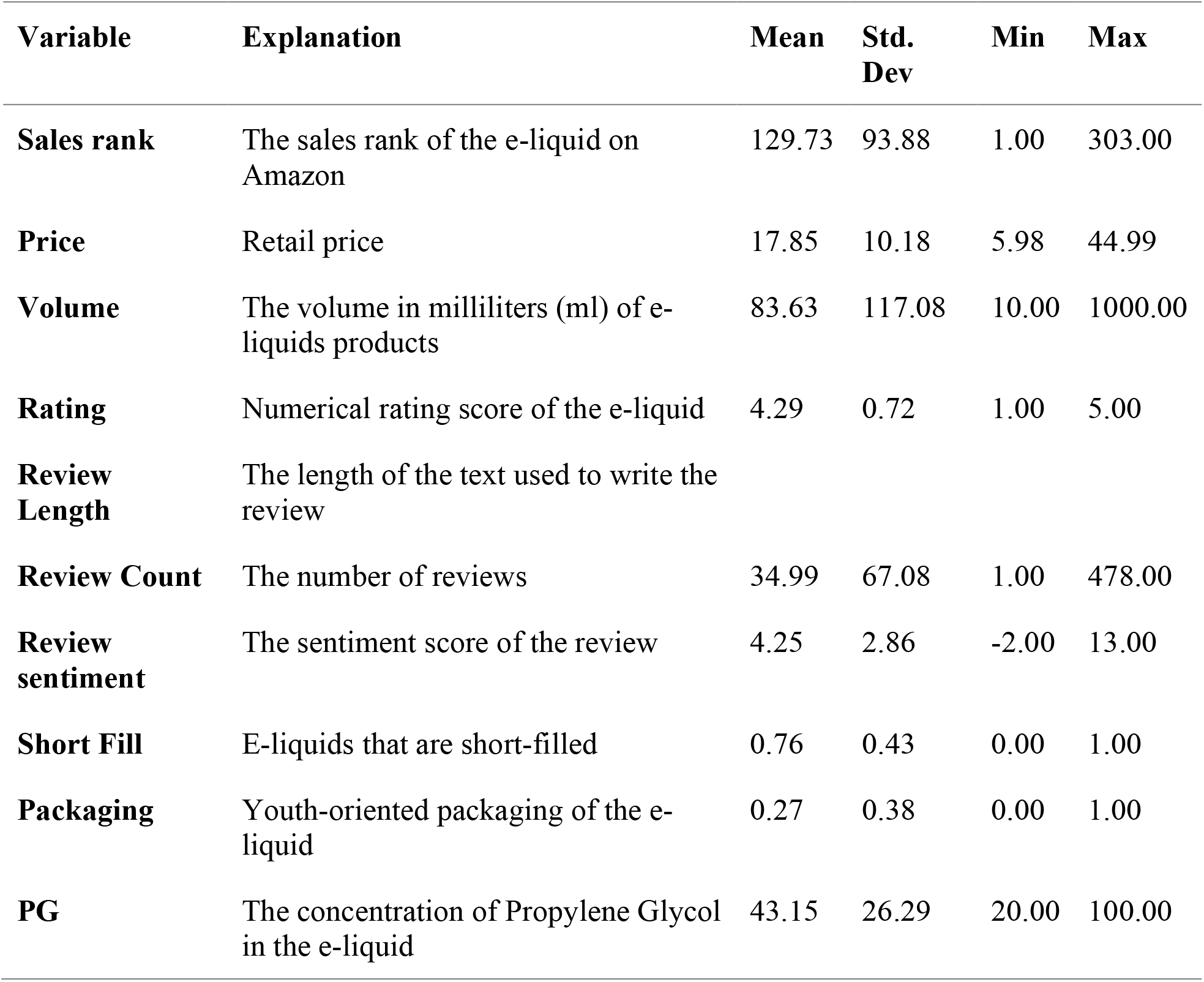
Descriptive statistics of e-liquids sold on Amazon in the United Kingdom.

### Empirical Results and discussion

We estimate our regression using the Ordinary Least Square (model1) and the mixed-effect linear regression model (model 2). We use the full maximum likelihood (Bates, et al., 2014, Hox, et al., 2017, Yang, et al., 2016) to produce robust, asymptotically efficient, and consistent estimates with the mixed linear models. Table 2 presents the results of the estimates.

**Table 2:** Estimation results of OLS and mixed-effect regression models.

| Variable | Model 1<br>OLS | Model 2<br>Mixed-effects |
| --- | --- | --- |
| Packaging | -0.74**<br>(0.30) | -0.58***<br>(0.18) |
| Sentiment | 0.08**<br>(0.03) | -0.01***<br>(0.00) |
| Log (Price) | -0.22<br>(0.20) | 0.02**<br>(0.01) |
| Log (Volume) | -0.64**<br>(0.30) | -0.07**<br>(0.04) |
| Log (Rating) | -0.15<br>(0.28) | -0.20***<br>(0.08) |
| Review Count | -0.18<br>(0.45) | -0.04*<br>(0.03) |
| Length Review | -0.001**<br>(0.00) | 0.00***<br>(0.00) |
| Short Fill | -0.92**<br>(0.38) | 4.15***<br>(0.61) |
| PG | -0.03***<br>(0.00) | -0.006**<br>(0.003) |
| Intercept | 10.14***<br>(1.87) | 2.26***<br>(0.43) |
| **Random effects** |  |  |
| Var($\mu$) | | 1.6955 |
| Var($v$) | | 0.0041 |
| Var($\epsilon$) | | 0.0005 |
| AIC | 178.58 | 54.52 |
| BIC | 201.80 | 138.95 |

(Notes: The dependent variable is Log (SalesRank); \*\*\*, \*\*, and \* indicate respectively significance at 0.01, 0.05, and 0.10 levels. Robust standard errors are presented in parentheses. Var( $\mu$ ), Var( $v$ ), Var( $\epsilon$ ) indicate respectively the variance of the brand-level random effect, the variance of the e-liquid random effect, and the variance of the error term.).

Based on the AIC and BIC criterion, the mixed-effects models perform betters than the OLS. The estimates of the random effects variances show that the brands explains 99% (1.6955/ (1.6955+0.0041+0.0005)) of the total variance. The OLS model ignores the hierarchical structure of the dataset, which leads to biased standard errors and inaccurate statistical significance. The rest of our analysis will be focused on the estimates derived from the mixed-effect linear regression model.

Our main interest is to evaluate the effect of using youth-oriented packaging on the demand for e-liquid. The negative sign and significance of the estimate for the Packaging variable suggest that e-liquid products with youth-oriented (kiddy and cartoonish) packaging sell better (lower sale rank) compared to e-liquid products with non-youth-oriented packaging. This result is consistent with previous findings on the effect of packaging on demand (Al‐Samarraie, et al., 2019, Fraser, 2018). This result can be explained by the fact that youth-oriented packaging might give Similar to previous studies that show that fruity flavors lead to an increase in e-cigarettes use (Ali, et al., 2020, Leventhal, et al., 2019, Wang, et al., 2020, Zare and Zheng, 2020), the negative and significant packaging covariate suggests that youth-enticing packaging will also induce a higher demand for e-liquids with the public health risks associated with their use (Eaton, et al., 2018). An implication for public health is that policies similar to the one related to the ban of youth-appealing flavors (including fruit and mint) in e-cigarettes should also be considered to regulate the packaging of e-liquids.

The coefficient on price is significantly different from zero and has a positive sign showing that the law of demand holds. As the price of e-liquid products increases, the quantity demanded by consumers decrease (higher sale rank). Using the estimated coefficient on the logarithm of price in table 2, and previous studies that show that sales rank can be used as a proxy for demand (Chevalier and Mayzlin, 2006, Forman, et al., 2008), the own-price elasticity of e-liquid demand can be estimated at 0.02. This shows that the demand for e-liquid is very inelastic on the Amazon platform. A 1% increase in the price of e-liquids will lead to a decrease in the quantity demanded by only 0.02%. the quantity demanded by e-liquid users is unresponsive to change in price which is consistent with Pope, et al. (2019). A major health policy question associated with the negative impact e-cigarettes consumption has on health, especially among the youth, is the extent to which tax policy can be used to reduce e-cigarette consumption. The efficacy of taxation depends on a large price elasticity of demand. Inelastic demand implies that policies such as taxes might not be effective on the demand for e-liquids.

The coefficient of the Volume variable is negative and statistically significant suggesting that larger e-liquid packages are associated with higher sales.

All four of the rating/review coefficients (rating, number of reviews or review count, average length of reviews, and sentiment in the reviews) estimated in our model are significantly different from zero. The negative coefficients on rating and review count indicate that an increase in the rating score and the number of reviews is associated with an increase in the sales of e-liquid. E-liquids with higher rating scores imply a positive evaluation of the performance or quality of the e-liquid by previous buyers, inducing the purchase decision. This result is consistent with the literature on the impact of rating for online platforms sales (Forman, et al., 2008, Ghose and Ipeirotis, 2010). Also, products that have been reviewed by more buyers have higher sales. The length of the review has a positive sign indicating that longer reviews do not increase sales. However, the estimate of the coefficient of the length of the review (0.00) suggests that this variable does not have a meaningful impact on sales.

Potential buyers form an expectation about the performance of the product by reading the reviews. Their purchasing decision depends on the expectation (positive or negative) derived from reading the reviews related to the product. This indicates that the content of the review plays an important role in sales. We investigated this hypothesis using sentiment analysis. Following Lawani, et al. (2019) we derived the sentiment in the reviews and examine its effect on sales. The coefficient on the sentiment variable in our econometric model is estimated to be negative and significant, which confirms the hypothesis that the sentiment in the reviews increase sales. A one-point increase in the sentiment score is associated with a one-point increase in the sales rank.

E-liquids with higher concentrations of propylene glycol (PG) sell better on the online platform as shown by the negative and significant coefficient. One possible explanation is the stronger throat hit sensation e-liquid with higher PG provides. Traditional cigarettes also provide a strong throat hit sensation. Smokers might substitute traditional cigarettes for e-cigarettes but use e-liquid with higher PG. However, higher concentrations of PG in e-liquid increase the risks of lung disease especially among youth (Choi, et al., 2010, Madison, et al., 2019). This is another worrisome result from the analysis.

Finally, and contrary to our expectation, short-fill e-liquids sell less than e-liquids that are not short-fill. Short-fill e-liquids offer the possibility for end-users to adjust the concentration of nicotine in their e-juice. This flexibility offered by short-fill e-liquids would seem to increase their sales. The positive and significant coefficient of the short-fill variable in our regression model suggests otherwise. A useful interpretation of this result is that buyers of e-liquids on Amazon expect a product ready to use instead of one that requires additional adjustments.

## Conclusion

Electronic nicotine delivery systems (ENDS) such as e-cigarettes pose a considerable public health threat to youth in many countries. The marketing of these products gives a no or low low-harm effect perception to their users. For example, the promotion of ENDS with different flavors such as fruit, candy, and mint diminishes the health hazards associated with these products and instead highlights possible pleasant experiences with these products making them attractive to youth. Regulatory institutions have issued policies that regulate fruit and mint flavors but have failed to address packaging, which can have similar enticing effects and drive youth to purchase, try-out, and keep using e-cigarettes. This failure might be because there is little scientific evidence on the effects of packaging on demand for e-cigarettes. Several studies have examined the effects of e-cigarettes attributes such as flavors and advertising on e-cigarettes sales. However, few studies examine the effect of packaging on purchases. No known study has explored the effect of youth-oriented packages on online e-liquid sales. This study contributes to the current literature by examining the impact of e-liquids packaging on demand and provides evidence for regulatory bodies to consider regulating ENDS packaging.

As online platforms are a significant means for e-liquid purchasing among the youth and reviews online influence purchase decisions, this study uses novel data scraped from Amazon to examine the effect of packaging on sales. Data on retail price, sales rank (used as a proxy for demand), packaging, and ratings/reviews were used in our analysis. The results indicate that e-liquids with youth-oriented packaging sell better. Online purchasers of a product often leave a review to encourage others to purchase or deter them from purchasing. The narrative for e-liquids is no different. Consistent with previous studies on the impact of reviews on sales (Chevalier and Mayzlin, 2006, Ghose and Ipeirotis, 2010), we find that higher rating scores translate to higher sales of e-liquids. Moreover, products with more reviews have higher sales. Positive sentiments in the reviews also appear to increase sales.

This study highlights how youth-oriented packaging of e-liquid products may be contributing as much to the increased demand for ENDS and thus to the ongoing pandemic of e-cigarettes use among the youth than initially examined. The findings of the study suggest that regulating youth-oriented packaging that typically targets the youth may help reduce purchases. Given the significant public health concerns associated with e-cigarette use among the youth, the question of whether to use taxes as a regulatory mechanism to curb their proliferation among youth persists. We find that the demand for e-liquids is price inelastic. This finding suggests that an e-liquid-tax may not be an effective policy tool for controlling demand.

Although this paper shows how youth-oriented packaging can influence the demand for e-liquids, future research should be conducted to expand the data from Amazon.co.uk to other countries in which Amazon operates as well as collect data from other online platforms that sell ENDS products.

## Data Availability

The data used in this research are available upon request

## Footnotes

1 On Amazon, e-liquids are also listed as e-juice or vape juice. We scrapped the data on the three keywords: e-liquids, e-juice, and vape-juice.

## Reference

Al-Samarraie, H., A. Eldenfria, J.E. Dodoo, A.I. Alzahrani, and N. Alalwan. 2019. “Packaging design elements and consumers’ decision to buy from the Web: A cause and effect decision-making model.” Color Research & Application 44:993–1005.

Ali, F.R.M., M.C. Diaz, D. Vallone, M.A. Tynan, J. Cordova, E.L. Seaman, K.F. Trivers, B.A. Schillo, B. Talley, and B.A. King. 2020. “E-cigarette Unit Sales, by Product and Flavor Type—United States, 2014–2020.” Morbidity and Mortality Weekly Report 69:1313.

Awan, K.H. 2016. “Experimentation and correlates of electronic nicotine delivery system (electronic cigarettes) among university students – A cross sectional study.” The Saudi Dental Journal 28:91–95.

Bates, D., M. Mächler, B. Bolker, and S. Walker. 2014. “Fitting linear mixed-effects models using lme4.” arXiv preprint 1406.5823.

Blount, B.C., M.P. Karwowski, P.G. Shields, M. Morel-Espinosa, L. Valentin-Blasini, M. Gardner, M. Braselton, C.R. Brosius, K.T. Caron, D. Chambers, J. Corstvet, E. Cowan, V.R. De Jesus, P. Espinosa, C. Fernandez, C. Holder, Z. Kuklenyik, J.D. Kusovschi, C. Newman, G.B. Reis, J. Rees, C. Reese, L. Silva, T. Seyler, M.A. Song, C. Sosnoff, C.R. Spitzer, D. Tevis, L. Wang, C. Watson, M.D. Wewers, B. Xia, D.T. Heitkemper, I. Ghinai, J. Layden, P. Briss, B.A. King, L.J. Delaney, C.M. Jones, G.T. Baldwin, A. Patel, D. Meaney-Delman, D. Rose, V. Krishnasamy, J.R. Barr, J. Thomas, J.L. Pirkle, and G. Lung Injury Response Laboratory Working. 2020. “Vitamin E Acetate in Bronchoalveolar-Lavage Fluid Associated with EVALI.” N Engl J Med 382:697–705.

Chen-Sankey, J.C., J.B. Unger, M. Bansal-Travers, J. Niederdeppe, E. Bernat, and K. Choi. 2019. “E-cigarette Marketing Exposure and Subsequent Experimentation Among Youth and Young Adults.” Pediatrics 144.

Chevalier, J., and A. Goolsbee. 2003. “Measuring prices and price competition online: Amazon. com and BarnesandNoble. com.” Quantitative marketing and Economics 1:203–222.

Chevalier, J.A., and D. Mayzlin. 2006. “The effect of word of mouth on sales: Online book reviews.” Journal of Marketing Research 43:345–354.

Choi, H., N. Schmidbauer, J. Sundell, M. Hasselgren, J. Spengler, and C.G. Bornehag. 2010. “Common household chemicals and the allergy risks in pre-school age children.” PLoS One 5:e13423.

Cole-Lewis, H., J. Pugatch, A. Sanders, A. Varghese, S. Posada, C. Yun, M. Schwarz, and E. Augustson. 2015. “Social Listening: A Content Analysis of E-Cigarette Discussions on Twitter.” Journal of Medical Internet Research 17:e243.

Collins, L., A.M. Glasser, H. Abudayyeh, J.L. Pearson, and A.C. Villanti. 2019. “E-Cigarette Marketing and Communication: How E-Cigarette Companies Market E-Cigarettes and the Public Engages with E-cigarette Information.” Nicotine Tob Res 21:14–24.

Conner, M., S. Grogan, R. Simms-Ellis, K. Flett, B. Sykes-Muskett, L. Cowap, R. Lawton, C.J. Armitage, D. Meads, C. Torgerson, R. West, and K. Siddiqi. 2018. “Do electronic cigarettes increase cigarette smoking in UK adolescents? Evidence from a 12-month prospective study.” Tobacco control 27:365–372.

Czoli, C.D., D. Hammond, and C.M. White. 2014. “Electronic cigarettes in Canada: Prevalence of use and perceptions among youth and young adults.” Canadian Journal of Public Health = Revue Canadienne de Santé Publique 105:e97–e102.

Eaton, D.L., L.Y. Kwan, and K. Stratton. 2018. “Public health consequences of e-cigarettes.”

El-Hage, R., A. El-Hellani, R. Salman, S. Talih, A. Shihadeh, and N.A. Saliba. 2020. “Vaped Humectants in E-Cigarettes Are a Source of Phenols.” Chem Res Toxicol 33:2374–2380.

Etter, J.F. 2018. “Gateway effects and electronic cigarettes.” Addiction 113:1776–1783.

Filippidis, F.T., A.A. Laverty, V. Gerovasili, and C.I. Vardavas. 2017. “Two-year trends and predictors of e-cigarette use in 27 European Union member states.” Tobacco control 26:98–104.

Forman, C., A. Ghose, and B. Wiesenfeld. 2008. “Examining the relationship between reviews and sales: The role of reviewer identity disclosure in electronic markets.” Information Systems Research 19:291–313.

Fraser, A. 2018. “The influence of package design on consumer purchase intent.” Packaging for Nonthermal Processing of Food:225–249.

Ghose, A., and P.G. Ipeirotis. 2010. “Estimating the helpfulness and economic impact of product reviews: Mining text and reviewer characteristics.” IEEE transactions on knowledge and data engineering 23:1498–1512.

Grace, R.C., B.M. Kivell, and M. Laugesen. 2014. “Estimating Cross-Price Elasticity of E-Cigarettes Using a Simulated Demand Procedure.” Nicotine & Tobacco Research 17:592–598.

Guo, X., H.-K. Lui, and G. Cui. 2012. “The Effect of Online Consumer Reviews on New Product Sales.” International Journal of Electronic Commerce 17:39–58.

Hamdar, B.C., M. Al Dana, and G. Al Chawa. 2018. “Economic Effects of Product Packaging on Consumer Shopping Behavior: The Case of Lebanon.” American Journal of Theoretical and Applied Business 4:44–47.

Hammond, D., O.A. Wackowski, J.L. Reid, R.J. O’Connor, and t. The International Tobacco Control Policy Evaluation Project. 2020. “Use of JUUL E-cigarettes Among Youth in the United States.” Nicotine & Tobacco Research 22:827–832.

Hox, J.J., M. Moerbeek, and R. Van de Schoot. 2017. Multilevel analysis: Techniques and applications: Routledge.

Hu, N., N.S. Koh, and S.K. Reddy. 2014. “Ratings lead you to the product, reviews help you clinch it? The mediating role of online review sentiments on product sales.” Decision Support Systems 57:42–53.

Hussain, S., S. Ali, M. Ibrahim, A. Noreen, and S. Ahmad. 2011. “Impact of product packaging on consumer perception and purchase intention.” Journal of Marketing and Consumer Research 10:1–10.

Kilcommons, S., S. Horwitz, S. Eon Ha, K. Ebbert, L. Restivo, M.-C.M. Verbeke, A. Hays-Alberstat, L. Cooke, C. Mackay, M. Anselmo, I. Mitchell, C.J. Doig, and J.R. Guichon. 2020. “Is Canadian federal legislation effective in preventing youth access to vaping initiation products? A study using secret shoppers and online access in three Alberta cities.” Preventive Medicine Reports 19:101117.

Kinouani, S., E. Pereira, and C. Tzourio. 2017. “Electronic Cigarette Use in Students and Its Relation with Tobacco-Smoking: A Cross-Sectional Analysis of the i-Share Study.” International Journal of Environmental Research and Public Health 14:1345.

Kong, G., M.E. Morean, D.A. Cavallo, D.R. Camenga, and S. Krishnan-Sarin. 2015. “Reasons for Electronic Cigarette Experimentation and Discontinuation Among Adolescents and Young Adults.” Nicotine & Tobacco Research 17:847–854.

Krishnasamy, V.P., B.D. Hallowell, J.Y. Ko, A. Board, K.P. Hartnett, P.P. Salvatore, M. Danielson, A. Kite-Powell, E. Twentyman, and L. Kim. 2020. “Update: characteristics of a nationwide outbreak of e-cigarette, or vaping, product use–associated lung injury— United States, August 2019–January 2020.” Morbidity and Mortality Weekly Report 69:90.

Kwon, M., and E. Park. 2020. “Perceptions and Sentiments About Electronic Cigarettes on Social Media Platforms: Systematic Review.” JMIR Public Health and Surveillance 6:e13673.

Lanza, H.I., A.M. Leventhal, J. Cho, J.L. Braymiller, E.A. Krueger, R. McConnell, and J.L. Barrington-Trimis. 2020. “Young adult e-cigarette use: A latent class analysis of device and flavor use, 2018-2019.” Drug and Alcohol Dependence 216:108258.

Lawani, A., M.R. Reed, T. Mark, and Y. Zheng. 2019. “Reviews and price on online platforms: Evidence from sentiment analysis of Airbnb reviews in Boston.” Regional Science and Urban Economics 75:22–34.

Lee, J.A., S. Lee, and H.-J. Cho. 2017. “The Relation between Frequency of E-Cigarette Use and Frequency and Intensity of Cigarette Smoking among South Korean Adolescents.” International Journal of Environmental Research and Public Health 14:305.

Leventhal, A.M., N.I. Goldenson, J. Cho, M.G. Kirkpatrick, R.S. McConnell, M.D. Stone, R.D. Pang, J. Audrain-McGovern, and J.L. Barrington-Trimis. 2019. “Flavored e-cigarette use and progression of vaping in adolescents.” Pediatrics 144.

Lu, X., L. Chen, J. Yuan, J. Luo, J. Luo, Z. Xie, and D. Li. 2020. “User Perceptions of Different Electronic Cigarette Flavors on Social Media: Observational Study.” Journal of Medical Internet Research 22:e17280.

MacDonald, A., and H.R. Middlekauff. 2019. “Electronic cigarettes and cardiovascular health: what do we know so far?” Vascular Health and Risk Management 15:159–174.

Madison, M.C., C.T. Landers, B.H. Gu, C.Y. Chang, H.Y. Tung, R. You, M.J. Hong, N. Baghaei, L.Z. Song, P. Porter, N. Putluri, R. Salas, B.E. Gilbert, I. Levental, M.J. Campen, D.B. Corry, and F. Kheradmand. 2019. “Electronic cigarettes disrupt lung lipid homeostasis and innate immunity independent of nicotine.” J Clin Invest 129:4290–4304.

Mantey, D.S., M.R. Cooper, S.L. Clendennen, K.E. Pasch, and C.L. Perry. 2016. “E-Cigarette Marketing Exposure Is Associated With E-Cigarette Use Among US Youth.” Journal of Adolescent Health 58:686–690.

Marynak, K., A. Gentzke, T.W. Wang, L. Neff, and B.A. King. 2018. “Exposure to Electronic Cigarette Advertising Among Middle and High School Students — United States, 2014– 2016.” Morbidity and Mortality Weekly Report 67:294–299.

McCausland, K., B. Maycock, T. Leaver, and J. Jancey. 2019. “The Messages Presented in Electronic Cigarette–Related Social Media Promotions and Discussion: Scoping Review.” Journal of Medical Internet Research 21:e11953.

McMillen, R.C., M.A. Gottlieb, R.M.W. Shaefer, J.P. Winickoff, and J.D. Klein. 2015. “Trends in Electronic Cigarette Use Among U.S. Adults: Use is Increasing in Both Smokers and Nonsmokers.” Nicotine & Tobacco Research 17:1195–1202.

Muhammad, A., and A. Kamran. 2014. “Impact of visual elements of packaging of packaged milk on consumer buying behaviour.” Interdiscip. J. Contemp. Res. Bus 5:118–160.

Nielsen, F.Å. 2011. “A new ANEW: Evaluation of a word list for sentiment analysis in microblogs.” arXiv preprint 1103.2903.

Orford, S. 2000. “Modelling spatial structures in local housing market dynamics: A multilevel perspective.” Urban Studies 37:1643–1671.

Pesko, M.F., J. Huang, L.D. Johnston, and F.J. Chaloupka. 2018. “E-cigarette price sensitivity among middle- and high-school students: evidence from monitoring the future.” Addiction 113:896–906.

Pope, D.A., L. Poe, J.S. Stein, B.A. Kaplan, W.B. DeHart, A.M. Mellis, B.W. Heckman, L.H. Epstein, F.J. Chaloupka, and W.K. Bickel. 2019. “The Experimental Tobacco Marketplace: Demand and Substitutability as a Function of Cigarette Taxes and e-Liquid Subsidies.” Nicotine & Tobacco Research 22:782–790.

Qasim, H., Z.A. Karim, J.O. Rivera, F.T. Khasawneh, and F.Z. Alshbool. 2017. “Impact of Electronic Cigarettes on the Cardiovascular System.” Journal of the American Heart Association 6.

Raheem, A.R., P. Vishnu, and A.M. Ahmed. 2014. “Impact of product packaging on consumer’s buying behavior.” European journal of scientific research 122:125–134.

Reid, J.L., V.L. Rynard, C.D. Czoli, and D. Hammond. 2015. “Who is using e-cigarettes in Canada? Nationally representative data on the prevalence of e-cigarette use among Canadians.” Preventive medicine 81:180–183.

Reinhold, B., R. Fischbein, S.S. Bhamidipalli, J. Bryant, and D.R. Kenne. 2017. “Associations of attitudes towards electronic cigarettes with advertisement exposure and social determinants: a cross sectional study.” Tobacco Induced Diseases 15:13.

Schneider, S., and K. Diehl. 2016. “Vaping as a Catalyst for Smoking? An Initial Model on the Initiation of Electronic Cigarette Use and the Transition to Tobacco Smoking Among Adolescents.” Nicotine Tob Res 18:647–653.

Struik, L.L., S. Dow-Fleisner, M. Belliveau, D. Thompson, and R. Janke. 2020. “Tactics for Drawing Youth to Vaping: Content Analysis of Electronic Cigarette Advertisements.” Journal of Medical Internet Research 22:e18943.

Tuchman, A.E. 2019. “Advertising and demand for addictive goods: The effects of e-cigarette advertising.” Marketing science 38:994–1022.

Tucker, M.R., M. Laugesen, C. Bullen, and R.C. Grace. 2017. “Predicting Short-Term Uptake of Electronic Cigarettes: Effects of Nicotine, Subjective Effects, and Simulated Demand.” Nicotine & Tobacco Research 20:1265–1271.

Venter, K., D. Van der Merwe, H. De Beer, E. Kempen, and M. Bosman. 2011. “Consumers’ perceptions of food packaging: an exploratory investigation in Potchefstroom, South Africa.” International Journal of Consumer Studies 35:273–281.

Walley, S.C., K.M. Wilson, J.P. Winickoff, and J. Groner. 2019. “A Public Health Crisis: Electronic Cigarettes, Vape, and JUUL.” Pediatrics 143.

Wang, T.W., L.J. Neff, E. Park-Lee, C. Ren, K.A. Cullen, and B.A. King. 2020. “E-cigarette use among middle and high school students—United States, 2020.” Morbidity and Mortality Weekly Report 69:1310.

Xiao, L., M. Parascandola, C. Wang, and Y. Jiang. 2018. “Perception and Current Use of E-cigarettes Among Youth in China.” Nicotine & Tobacco Research 21:1401–1407.

Yang, Y., N.J. Mueller, and R.R. Croes. 2016. “Market accessibility and hotel prices in the Caribbean: The moderating effect of quality-signaling factors.” Tourism Management 56:40–51.

Zare, S., and Y. Zheng. 2020. “Consumer Preferences for E-cigarette Flavor, Nicotine Strength, and Type: Evidence from Nielsen Scanner Data.” Nicotine Tob Res.

